# Clinical Implications of Atrial Fibrillation Provoked by Acetylcholine

**DOI:** 10.1101/2023.12.27.23300593

**Authors:** Keita Shibata, Kohei Wakabayashi, Naoko Ikeda, Tomoyuki Ishinaga, Yuta Kusakabe, Masaki Asakawa, Naoki Aizawa, Suguru Shimazu, Takahiro Furuya, Yuya Nakamura, Chisato Sato, Tenjin Nishikura, Masaru Shiigai, Mitsunori Mutou, Junko Honye, Kaoru Tanno

## Abstract

**Background:** The coronary spasm provocation test using acetylcholine (ACh) is useful for diagnosing vasospastic angina (VSA). Paroxysmal atrial fibrillation (PAF) during ACh testing is frequent and usually transient. However, the clinical implications of PAF provoked by ACh is unknown. Deterioration of the left atrial (LA) reservoir strain is associated with new-onset of atrial fibrillation (AF) and fibrosis of the left atrium; additionally, ACh shortens the action potential duration and facilitates AF in the fibrotic atria in the human AF model. Hence, this study aimed to investigate the relationship between LA function and occurrence of AF during the ACh test.

**Methods:** We studied a consecutive cohort of 100 patients (60.1±14.5 years, 39 women) without history of AF who underwent the ACh test in our centers from 2015 to 2022. Echocardiographic data were available for all the patients. PAF was defined as lasting >30 s during the ACh test. Based on the occurrence of AF during the ACh test, the patients were divided into two groups: provoked PAF group (n=29) vs. non AF group (n=71). LA function was assessed via two-dimensional speckle-tracking echocardiography. Occurrences of spontaneous AF were recorded as clinical events during long-term follow-up.

**Results:** The observation period was 675 (114.5-1789.5) days, and 65 patients (65%) were tested positive in the ACh test and diagnosed with VSA. LA volume index was similar between provoked PAF and non AF groups (26.9±7.4 mL/m^2^ vs. 27.1±8.5 mL/m^2^, *p*=0.89). In contrast, LA reservoir (27.6±5.2% vs. 34.8±6.8%, *p*<0.001) and conduit (13.4±5.1% vs. 18.4±6.2%, *p*<0.001) strain were significantly lower in provoked PAF group than in non AF group. The provoked PAF group had a lower LA booster strain (14.1±5.6% vs. 16.4±6.3%, *p*=0.093) than non AF group. A multiple regression analysis showed that LA reservoir strain was independently associated with the provoked PAF during ACh test (OR 0.81, 95% CI: 0.72-0.91, *p*<0.001). In the provoked PAF group, spontaneous AF occurred in three patients (10.3%), one of whom was treated with pulmonary vein isolation, whereas there were no events of PAF in the non AF group.

**Conclusion:** The occurrence of PAF during the ACh test suggests dysfunction of left atria and may predict future PAF.

## Introduction

Coronary spasm provocation test using acetylcholine (ACh) is the gold standard for diagnosing vasospastic angina (VSA). [1–5] During the ACh test, paroxysmal atrial fibrillation (PAF) is frequently (8.0-22.8%) provoked, is usually transient, and sometimes requires treatment with antiarrhythmic drugs or electrical cardioversion. [6–10] Although PAF during the ACh test is generally recognized as a minor complication, it may have clinical implications.

Left atrial (LA) diameter enlargement, measured using transthoracic echocardiography, is associated with the incidence and recurrence of atrial fibrillation (AF). [11] The deterioration of the LA reservoir strain assessed via two-dimensional (2-D) speckle tracking echocardiography often precedes structural changes, such as LA enlargement, resulting in clinical AF events. [12–13] Furthermore, an impaired LA reservoir strain is associated with fibrosis of the left atria. [14–15] ACh reportedly shortens action potential duration and facilitates AF in the fibrotic atria of a human AF model. [16] The association between AF provoked by the ACh test and LA function remains unknown.

We hypothesized that patients with PAF provoked during the ACh test would have lower LA function because of fibrotic left atria. Therefore, we aimed to identify the relationship between LA function assessed using 2-D speckle tracking echocardiography and the occurrence of AF during the ACh test and to investigate the impact of PAF triggered during the ACh test on clinical outcomes.

## Methods

### Study Population and Design

We included consecutive patients who underwent the ACh test for diagnosing VSA between January 2015 and January 2022 at two institutes—Showa University Koto-Toyosu Hospital and Kikuna Memorial Hospital. Exclusion criteria were as follows: patients with a history of AF i.e. stage 3 or 4, as per the new proposed classification by the American College of Cardiology and American Heart Association Joint Committee on clinical practice guidelines, before the ACh test and those whose echocardiographic data were inadequate for analysis or were unavailable. [17] Provoked PAF was defined as PAF lasting >30 s during the ACh test. In accordance with the occurrence of PAF during the ACh test, the patients were divided into two groups: the provoked PAF group vs. non AF group.

To determine the association between PAF provoked during the ACh test and the subsequent development of spontaneous AF attacks, we recorded the occurrence of AF as a clinical endpoint. The other endpoints were all-cause mortality, stroke, and hospitalization for heart failure.

Informed consent for data use in the present study was obtained using an opt-out form. The study was conducted in accordance with the Declaration of Helsinki and approved by the ethics committee of Showa University.

Coronary angiography (CAG) was performed after discontinuing the use of medications, such as long-acting nitroglycerin, nicorandil, and calcium channel blockers, for at least 48 h. A temporary pacemaker was inserted in the right ventricular apex through the internal jugular, femoral, or brachial veins at a rate of 40 beats/min. A control CAG of the left coronary artery (LCA) and right coronary artery (RCA) was performed without nitroglycerine or isosorbide dinitrate. ACh was injected in incremental doses of 20 and 50 μg into the RCA and 50 and 100 μg into the LCA over a period of 20 s. CAG was performed with at least 1 min interval between each injection. Positive ACh test results were defined as transient total occlusion or focal 90% stenosis of the coronary artery with signs or symptoms of myocardial ischemia (angina pain and ischemic electrocardiogram change) or 90% diffuse vasoconstriction induced in two or more contiguous segments of the coronary artery. [3] CAG was then performed with a coronary injection of at least 1 mg of isosorbide dinitrate for each coronary artery.

### Conventional Echocardiography

Standard 2-D and Doppler measurements were performed in all patients according to the guidelines of the Joint American Society of Echocardiology and European Association of Cardiovascular Imaging. [18–19] Both left ventricular and LA volumes were measured using the biplane Simpson method, and LA volume was indexed per the body surface area. In the diastolic phase, the peak early diastolic velocity (E) and peak atrial filling velocity (A) were measured. The peak early diastolic (e’) mitral annular tissue motion velocities were measured with a sample volume placed at the septal and lateral side of mitral annular levels from 4-chamber views and averaged to obtain the mean e’. E velocity was indexed by mean e’ to obtain E/e’. The tricuspid regurgitant pressure gradient (TRPG) was measured from the maximal systolic tricuspid regurgitation jet velocity using a modified Bernoulli’s equation. The longitudinal deformation variable LA was measured using 2-D speckle-tracking echocardiography, according to the current consensus document of the Joint European Association of Cardiovascular Imaging and American Society of Echocardiology. [20] The apical 4-chamber was stored offline for at least three consecutive cycles at a frame rate of 40-80 frames/s. Strain analysis was performed using vendor-independent 2-D strain software (TOMTEC Imaging Systems, Munich, Germany). The LA endocardial border was manually traced to define the region of interest, excluding the pulmonary vein and LA appendage. The focus was positioned at an intermediate depth. Each parameter of the LA reservoir, LA conduit, and LA booster in the cardiac cycle were then calculated. The left ventricular end diastole (mitral valve closure) was set to zero to calculate LA strains. Each LA strain was defined as follows; LA reservoir strain was the peak value at the onset of LV filling, and LA booster strain was the peak value at the onset of atrial contraction. LA conduit strain was defined as the difference between the LA reservoir strain and LA booster strain. LA strain data was carefully reviewed by two expert cardiologists who were blinded to the clinical information.

### Statistical Analysis

Categorical variables are presented as frequencies and percentages and were compared between groups using the chi-squared or Fisher’s exact test, as appropriate. Continuous variables are presented as the mean ± standard deviation (SD) or median and interquartile range (IQR) for skewed data and compared between groups using 2-tailed, unpaired t-tests or, if parameters were not normally distributed, using the Mann–Whitney U-test. Logistic regression analysis was used to examine the predictors of PAF provoked during the ACh test. Odds ratios (OR) and 95% confidence interval (CI) for each covariate were calculated. In the multivariable logistic regression analysis, covariates (*p*<0.10) in the univariable analysis were included. The incidence of spontaneous AF attacks during the long-term follow-up was estimated utilizing the Kaplan–Meier method, and the difference between the two groups was assessed using a log-rank test. Statistical significance was set at *p*<0.05. Statistical analyses were performed using the JMP Pro version 16.0 software (SAS Institute Inc., Cary, NC, USA).

## Results

### Patient clinical characteristics

A total of 144 consecutive patients underwent the ACh test between January 2015 and January 2022 (Figure 1). Patients with a history of AF before the ACh (n=34) test and those whose echocardiographic data (n=10) were inadequate for analysis were excluded. One hundred patients who underwent the Ach test were analyzed, of which 65 (65%) tested positive in the ACh test and were diagnosed with VSA. A provoked PAF was detected in 29 patients during the ACh test. Accordingly, the patients were divided into two groups: provoked PAF (n=29) and non AF (n=71). The clinical characteristics of each group are presented in Table 1. Serum B-type natriuretic peptide levels tended to be higher in the provoked-PAF group than in the non AF group.

**Figure 1.**
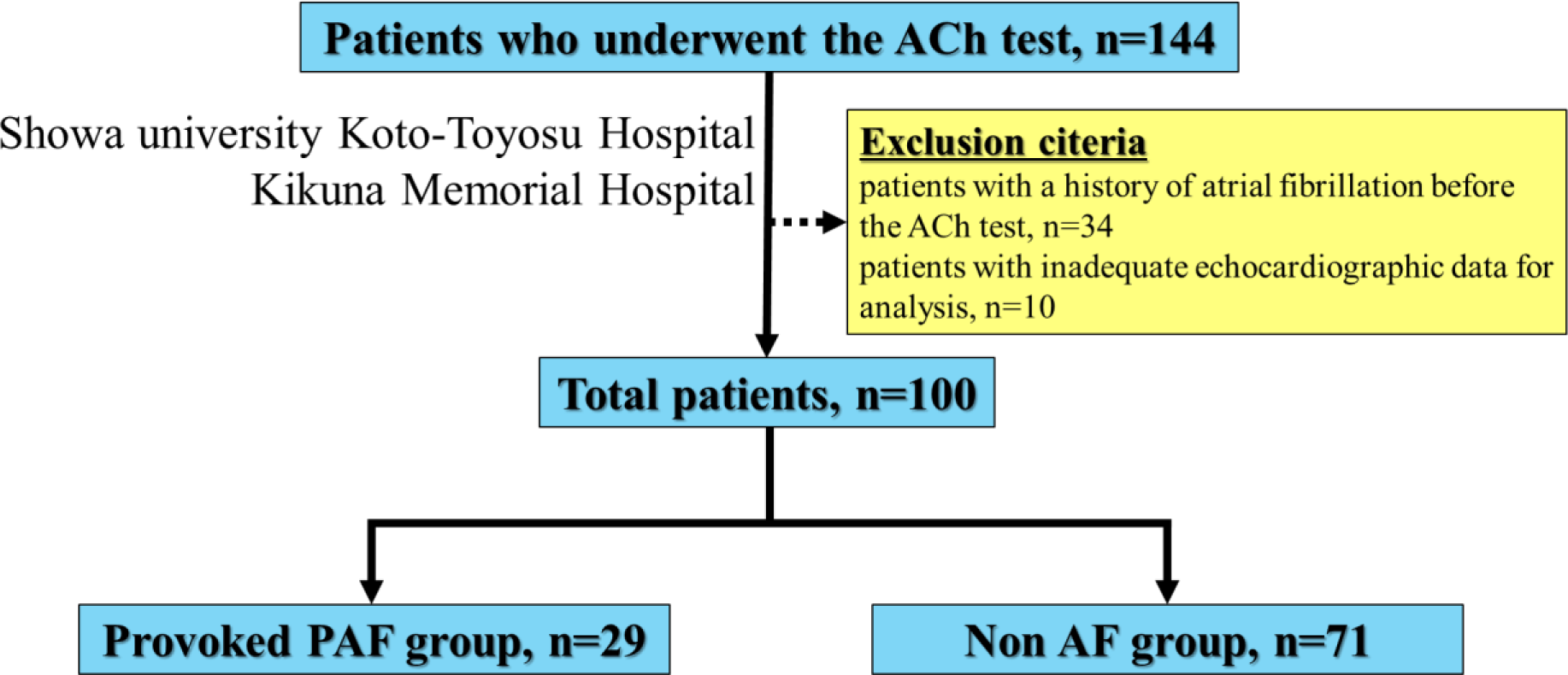
A total of 144 patients underwent ACh test at our center between January 2015 and January 2022. After excluding patients with a history of AF before the ACh test and those whose echocardiographic data were inadequate for analysis, the study comprised 100 patients. Based on the occurrence of AF during the ACh test, the patients were divided into two groups: the Provoked PAF group (n=29) vs. non AF group (n=71). ACh, acetylcholine; PAF, paroxysmal atrial fibrillation; AF, atrial fibrillation.

**Table 1.** Patient characteristics.

| Characteristics | Provoked PAF group<br>(n=29) | Non AF group<br>(n=71) | *p* value |
| --- | --- | --- | --- |
| Age, year | 61.5±13.9 | 58.6±13.8 | 0.35 |
| Female sex | 27 (38.0) | 12 (41.3) | 0.75 |
| Body mass index, kg/cm<sup>2</sup> | 23.8±5.0 | 24.3±3.7 | 0.66 |
| Vasospastic angina | 45 (63.3) | 20 (68.9) | 0.59 |
| Hypertension | 13 (44.8) | 33 (46.4) | 0.88 |
| Dyslipidemia | 14 (48.2) | 44 (61.9) | 0.20 |
| Diabetes mellitus | 7 (24.1) | 10 (14.0) | 0.22 |
| History of smoking | 14 (48.2) | 38 (53.5) | 0.63 |
| COPD | 0 (0) | 4 (5.6) | 0.32 |
| CKD | 2 (6.9) | 8 (11.2) | 0.71 |
| History of heart failure | 1 (3.4) | 1 (1.4) | 0.49 |
| Prior myocardial infarction | 0 (0) | 2 (2.8) | 1.00 |
| History of cerebrovascular disease | 1 (3.4) | 4 (5.6) | 1.00 |
| Laboratory data |  |  |  |
| Hemoglobin, g/mL | 14.0±1.4 | 13.9±1.2 | 0.78 |
| D dimer, µg/mL | 0.80±0.52 | 0.71±0.32 | 0.38 |
| Creatinine, mg/dL | 0.75±0.12 | 0.87±0.07 | 0.41 |
| eGFR, mL/min/1.73 m<sup>2</sup> | 76.8±17.5 | 72.7±17.6 | 0.29 |
| BNP, ng/mL | 11.8 (6.0-24.7) | 17.9 (8.0-41.9) | 0.12 |
| Antiarrhythmic drug at discharge |  |  |  |
| β blocker | 4 (13.7) | 5 (7.0) | 0.44 |
| Diltiazem | 4 (13.7) | 10 (14.0) | 1.00 |

Values are presented as n (%), mean±SD, and/or median (Q1-Q3). PAF, paroxysmal atrial fibrillation; AF, atrial fibrillation; COPD, chronic obstructive pulmonary disease; CKD, chronic kidney disease; eGFR, estimated glomerular filtration rate; BNP, B-type natriuretic peptide.

In transthoracic echocardiography, TRPG levels were significantly higher in Provoked PAF group (*p*=0.015). No significant difference in the left atrial dimension and left atrial volume index were noted between the groups, whereas the LA reservoir and conduit strains were significantly lower in the provoked PAF group than in the non AF group (*p*<0.001) (Table 2).

**Table 2.** Characteristics of transthoracic echocardiography.

| Transthoracic echocardiographic characteristics | Provoked PAF group (n=29) | Non AF group (n=71) | *p* value |
| --- | --- | --- | --- |
| LVEF, % | 63.2±1.3 | 62.2±0.8 | 0.51 |
| LVDd, mm | 46.3±4.7 | 46.3±5.5 | 0.99 |
| LVDs, mm | 29.7±5.4 | 30.3±5.4 | 0.60 |
| LAD, mL | 33.3±5.8 | 33.5±5.3 | 0.85 |
| LAVI, mL/m<sup>2</sup> | 26.9±7.4 | 27.1±8.5 | 0.89 |
| RA area, cm<sup>2</sup> | 11.8±2.3 | 12.4±3.0 | 0.34 |
| E wave velocity, cm/sec | 66.5±13.0 | 66.7±15.1 | 0.95 |
| A wave velocity, cm/sec | 72.1±23.9 | 69.5±23.0 | 0.62 |
| Mean e', cm/sec | 8.6±2.8 | 8.2±2.6 | 0.50 |
| E/e' | 9.3±3.4 | 9.2±2.9 | 0.86 |
| TRPG, mmHg | 20.9±5.8 | 17.7±5.7 | 0.015 |
| Estimated RVSP, mmHg | 24.5±6.5 | 21.4±6.3 | 0.036 |
| LA reservoir strain, % | 27.6±5.2 | 34.8±6.8 | <0.001 |
| LA booster strain, % | 13.4±5.1 | 18.4±6.2 | <0.001 |
| LA conduit strain, % | 14.1±5.6 | 16.4±6.3 | 0.093 |

Values are presented as n (%) and mean±SD. PAF, paroxysmal atrial fibrillation; AF, atrial fibrillation; LVEF, left ventricular ejection fraction; LVDd, left ventricular end-diastolic diameter; LVDs, left ventricular end-systolic diameter; LAD, left atrial dimension; LAVI, left atrial volume index; RA, right atrium; TRPG, tricuspid regurgitant pressure gradient; RVSP, right ventricular systolic pressure; LA, left atrial.

### Predictors of PAF provoked during the ACh test

There was a significant difference in LA reservoir and LA conduit strains between the provoked PAF and non-AF group (*p*<0.001), respectively (Figure 2). Univariable logistic regression analysis revealed that LA reservoir strain, LA conduit strain, and TRPG were the significant predictors of PAF provoked during the ACh test (Table 3). Instead of LA conduit strain, which is the difference between LA reservoir strain and LA booster strain, two variables, LA reservoir and LA booster strains, were included in the multivariable logistic analysis. After multivariable logistic regression analysis, lower LA reservoir strain was an independent predictor of PAF provoked during the ACh test (OR, 0.81; 95%CI, 0.73-0.91).

**Figure 2.**
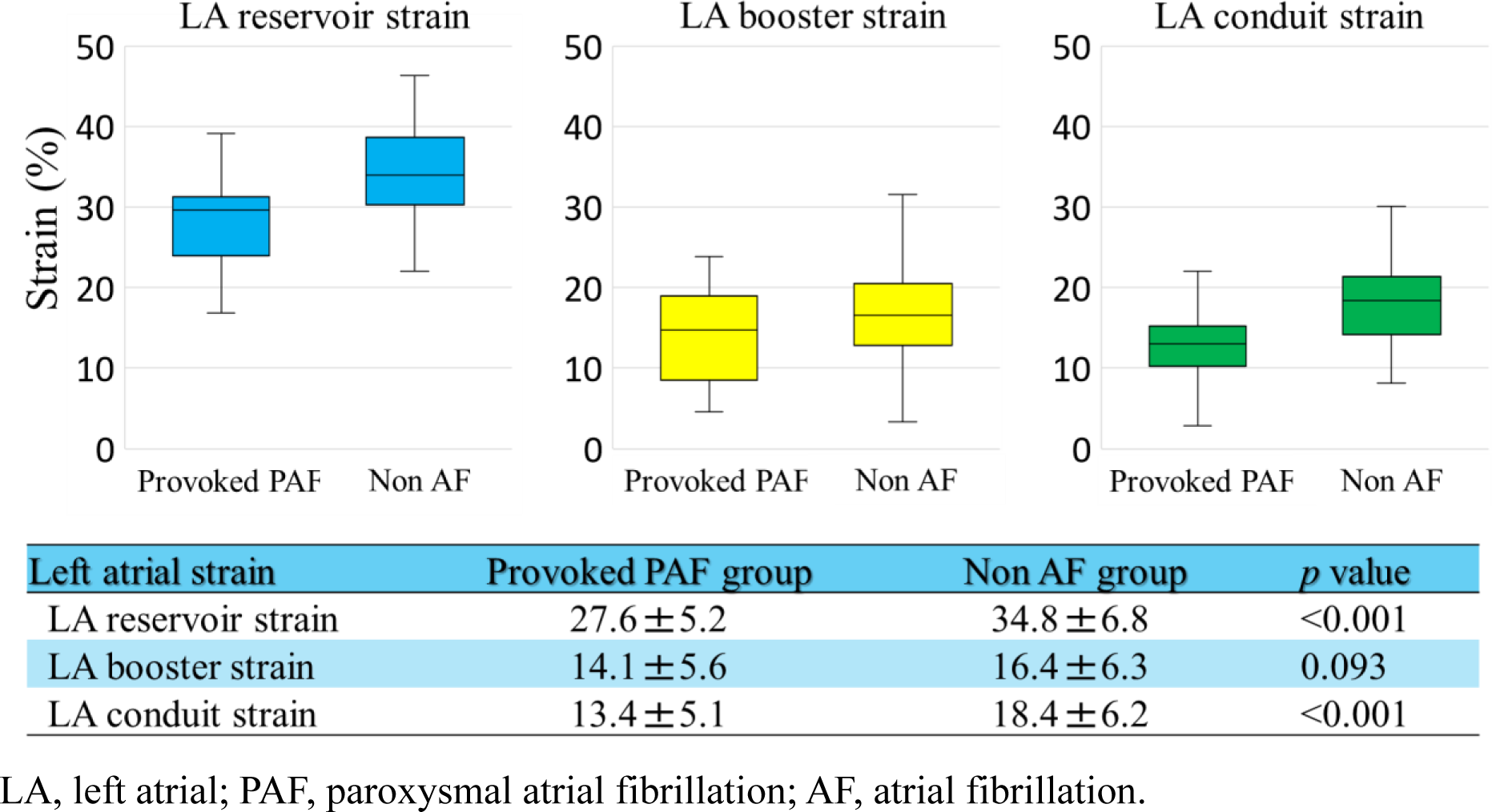
There was a significant difference in LA reservoir strain and LA conduit strain between the provoked PAF and non AF group. LA conduit strain is the difference between LA reservoir strain and LA booster strain. LA, left atrial; PAF, paroxysmal atrial fibrillation; AF, atrial fibrillation.

**Table 3.** Multivariable logistic regression for occurrence of provoked PAF during the Ach test.

|  | Univariable logistic analysis |  |  | Multivariable logistic analysis |  |  |
| --- | --- | --- | --- | --- | --- | --- |
|  | OR | (95% CI) | *p* value | OR | (95% CI) | *p* value |
| Age, year | 0.98 | (0.95-1.01) | 0.34 |  |  |  |
| Female sex | 1.15 | (0.47-2.77) | 0.75 |  |  |  |
| Body mass index, kg/cm<sup>2</sup> | 1.01 | (0.93-1.11) | 0.66 |  |  |  |
| Vasospastic angina | 1.28 | (0.51-3.23) | 0.59 |  |  |  |
| Hypertension | 0.93 | (0.39-2.22) | 0.88 |  |  |  |
| Dyslipidemia | 0.57 | (0.23-1.36) | 0.21 |  |  |  |
| Diabetes mellitus | 1.94 | (0.65-5.72) | 0.22 |  |  |  |
| History of smoking | 0.81 | (0.34-1.92) | 0.63 |  |  |  |
| CKD | 0.58 | (0.11-2.92) | 0.51 |  |  |  |
| BNP, pg/mL | 0.98 | (0.97-1.00) | 0.20 |  |  |  |
| TRPG, mmHg | 1.10 | (1.01-1.20) | 0.019 | 1.08 | (0.98-1.19) | 0.091 |
| LA reservoir strain, % | 0.80 | (0.72-0.89) | <0.001 | 0.81 | (0.72-0.91) | <0.001 |
| LA booster strain, % | 0.93 | (0.87-1.01) | 0.095 | 1.02 | (0.90-1.16) | 0.65 |
| (LA conduit strain, %) | 0.84 | (0.76-0.93) | <0.001 |  |  |  |

PAF, paroxysmal atrial fibrillation; AF, atrial fibrillation; OR, odds ratio; CI, confidence
interval; COPD, chronic obstructive pulmonary disease; CKD, chronic kidney disease; LA,
left atrial; TRPG, tricuspid regurgitant pressure gradient.

### Clinical outcomes

Clinical outcomes are summarized in Table 3. The median follow-up period was 575 (114.5-1789.5) days in the provoked PAF group and 675 (120-1644) days in the non AF group. In the provoked PAF group, three patients (10.3%) experienced spontaneous AF, one of whom was treated with catheter ablation, whereas there was no event of spontaneous AF in the non AF group. There was a significant difference in the Kaplan–Meier curves for PAF-free survival between the two groups (log-rank test, *p*=0.006; Figure 3). Stroke events and hospitalizations for heart failure have not been documented. During the follow-up period, two patients in the non PAF group experienced sudden cardiac death because of an uncontrolled VSA attack.

**Figure 3.**
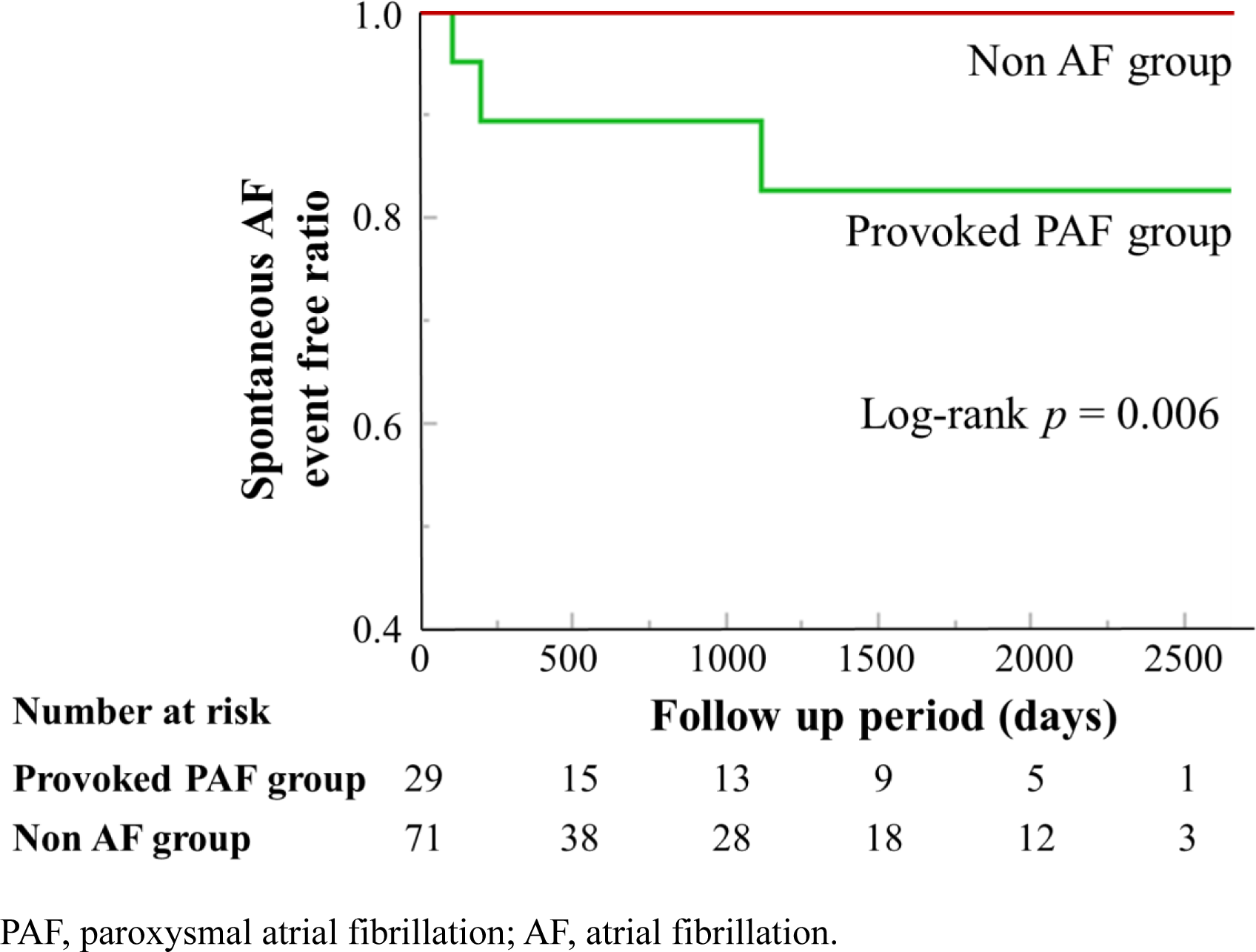
The median follow-up period was 575 (114.5–1789.5) days in the provoked PAF group and 675 (120–1644) days in the non AF group. In provoked PAF group, three patients (10.3%) experienced spontaneous AF, whereas there were no events of spontaneous AF in the non AF group. There was significant difference in the Kaplan–Meier curves for PAF-free survival between the two groups. AF atrial fibrillation; PAF paroxysmal atrial fibrillation.

**Table 4.** Clinical outcomes.

|  | Provoked PAF group<br>(n=29) | Non AF group<br>(n=71) | *p* value |
| --- | --- | --- | --- |
| Follow-up period, day | 575 (114.5-1789.5) | 675 (120-1644) | 0.32 |
| Occurrence of AF event | 3 (14.2) | 0 (0) | 0.022 |
| Pulmonary vein isolation | 1 (4.7) | 0 (0) | 0.29 |
| Stroke | 0 (0) | 0 (0) | 1.0 |
| Death | 0 (0) | 2 (2.8) | 1.0 |
| Heart failure | 0 (0) | 0 (0) | 1.0 |

Values are presented as n (%) and mean±SD. PAF, paroxysmal atrial fibrillation; AF, atrial fibrillation.

## Discussion

This study evaluated the relationship between PAF provoked during the ACh test and LA function and its clinical impact on long-term outcomes. The main findings of the present study were as follows: (1) patients with PAF provoked during the ACh test had significantly decreased LA reservoir strain, LA conduit strain, and higher TRPG; (2) deterioration of LA reservoir strain was an independent predictor of provoked PAF during the ACh test; and (3) during the follow-up period, spontaneous AF was observed only in patients with PAF provoked during the ACh test. The provocation test for coronary spasm using ACh is a general examination for the diagnosis of VSA. The triggering of PAF is a frequent complication associated with the ACh test. However, its clinical significance has been overlooked. To the best of our knowledge, this is the first report to assess the relationship between PAF provoked during the ACh test and LA function and its clinical implication.

### Relationship between left atrium dysfunction and PAF provoked by ACh

In atrial myocytes, the parasympathetic neurotransmitter ACh rapidly increases the outward potassium current (I_KACh_), and this reaction substantially shortens the action potential duration. [21] The association between AF and fibrosis in the left atrium has been systematically studied. [22–23] Previous reports have shown a close association between LA reservoir strain deterioration and PAF, which is associated with structural and mechanical remodeling in the left atrium. [12] Although LA fibrosis cannot be assessed directly on an echocardiogram, LA reservoir strain evaluated using 2-D speckle tracking echocardiography is associated with the extent of histologically confirmed LA fibrosis. [14–15]

In the present study, the deterioration of the LA reservoir strain was associated with PAF provoked during the ACh test. Although it is generally recognized that provoked AF during the ACh test is different from spontaneous AF, the results of the present study revealed a deterioration in LA reservoir strain even in provoked AF during the ACh test. Interestingly, the TRPG was higher in the provoked PAF group than in the non-AF group. It is conceivable that higher TRPG levels reflect an increase in the right ventricular pressure upstream of the fibrotic left atrium.

### Mechanism of provoked AF by ACh

Deterioration of LA reservoir strain is related to LA fibrosis, as mentioned above. Bayer et al. used human AF computer models composed of a LA appendage and the left atrium to study the effects of fibrosis on arrhythmogenicity using ACh. In fibrotic atria with heterogeneous parasympathetic activation, ACh facilitates AF by shortening the duration of the action potential and slowing conduction to promote a unidirectional conduction block and reentry. [16] Although this theory is based on an experimental model, it is possible that AF is provoked in patients exhibiting a decline in LA reservoir strain following ACh administration.

### Clinical implication of provoked PAF during the ACh test

It is believed that PAF provoked during the ACh test is minor and safe complication. In this study, spontaneous AF was observed only in the provoked PAF group during the ACh test. In the general population, LA reservoir strain (<23%) has been reported as a risk factor for future occurrence of spontaneous AF. [24] Additionally, in asymptomatic participants aged >65 years with more than one AF risk factor, including hypertension, diabetes mellitus, and obesity, an LA reservoir strain of less than 35% was independently associated with AF. [25] In the present study, the mean LA reservoir strain was 27.6% in the provoked PAF group. The results of the present study suggest that PAF provoked during the ACh test indicates potential fibrosis of left atria and predicts future spontaneous AF. The ACh test is the gold standard tool for diagnosing VSA, but it might also be useful for detecting potential deterioration of the LA reservoir and predicting the future occurrence of spontaneous AF. Further research is needed to confirm the results of the present study and to develop a novel test using ACh to determine the future risk of AF.

### Study limitations

This study had several limitations. First, the small sample size and relatively few spontaneous AF events may have influenced the results of our analysis. Second, it was impossible to detect all events of spontaneous AF events in patients in a long-term period. Thus, the occurrence of spontaneous AF may have been underestimated. Third, as we excluded patients with a history of AF, it is possible that some enrolled patients had undetected PAF. Finally, all the participants in this study were Japanese. Therefore, it is unclear whether these results can be generalized to other populations. Further international, large-scale studies are required to address this issue.

## Conclusion

Patients with PAF provoked by ACh more frequently exhibited deterioration of the LA reservoir strain. Spontaneous AF was observed only in patients with PAF provoked by the ACh test. This suggests that provoked PAF during the ACh test indicates potential fibrosis of the left atrium and the risk of future spontaneous AF.

## Data Availability

The data underlying this article are available in the article.

## Nonstandard Abbreviations and Acronyms

Ach: acetylcholine
AF: atrial fibrillation
BNP: B-type natriuretic peptide
CAG: coronary angiography
CKD: chronic kidney disease
CI: confidence interval
COPD: chronic obstructive pulmonary disease
eGFR: estimated glomerular filtration rate
IQR: interquartile range
LA: left atrial
LAD: left atrial dimension
LAV: left atrial volume
LAVI: left atrial volume index
LCA: left coronary artery
LVDd: left ventricular end-diastolic diameter
LVDs: left ventricular end-systolic diameter
LVEF: left ventricular ejection fraction
OR: odds ratio
PAF: paroxysmal atrial fibrillation
RA: right atrium
RCA: right coronary artery
RVSP: right ventricular systolic pressure
SD: standard deviation
TRPG: tricuspid regurgitant pressure gradient
VSA: vasospastic angina
2-D: two-dimensional

## Acknowledgments

The authors are grateful for the support of all staff at the Ultrasound Examination Center and Catheterization Laboratory at Showa University Koto-Toyosu Hospital and Kikuna Memorial Hospital.

